# Derivation and external validation of a clinical prognostic model identifying children at risk of death following presentation for diarrheal care

**DOI:** 10.1101/2023.02.08.23285625

**Authors:** Sharia M. Ahmed, Ben J. Brintz, Alison Talbert, Moses Ngari, Patricia B. Pavlinac, James A. Platts-Mills, Adam C. Levine, Eric J. Nelson, Judd L. Walson, Karen L. Kotloff, James A. Berkley, Daniel T. Leung

## Abstract

Diarrhea continues to be a leading cause of death for children under-five. Amongst children treated for acute diarrhea, mortality risk remains elevated during and after acute medical management. Identification of those at highest risk would enable better targeting of interventions, but available prognostic tools lack validation. We used clinical and demographic data from the Global Enteric Multicenter Study (GEMS) to build predictive models for death (in-treatment, after discharge, or either) in children aged ≤59 months presenting with moderate-to-severe diarrhea (MSD), in Africa and Asia. We screened variables using random forests, and assessed predictive performance with random forest regression and logistic regression using repeated cross-validation. We used data from the Kilifi Health and Demographic Surveillance System (KHDSS) and Kilifi County Hospital (KCH) in Kenya to externally validate our GEMS-derived clinical prognostic model (CPM). Of 8060 MSD cases, 43 (0.5%) children died in treatment and 122 (1.5% of remaining) died after discharge. MUAC at presentation, respiratory rate, age, temperature, number of days with diarrhea at presentation, number of people living in household, number of children <60 months old living in household, and how much the child had been offered to drink since diarrhea started were predictive of death both in treatment and after discharge. Using a parsimonious 2-variable prediction model, we achieve an AUC=0.84 (95% CI: 0.82, 0.86) in the derivation dataset, and an AUC=0.74 (95% CI 0.71, 0.77) in the external dataset. Our findings suggest it is possible to identify children most likely to die after presenting to care for acute diarrhea. This could represent a novel and cost-effective way to target resources for the prevention of childhood mortality.

## INTRO

Close to 500,000 children under 5 years of age die from diarrhea every year, mostly in low- and middle-income countries (LMICs). While children at higher risk of severe outcomes are more likely to be admitted to treatment[1], there is also growing recognition that the risk of death remains elevated even after treatment discharge[1-5]. In fact, some evidence suggests that young children may be at the greatest risk of death after being discharged from care[6, 7]. Clinicians may benefit from tools to help identify children at greater risk of death, in order to target them for additional care or follow-up interventions[5, 6].

In this study, we aimed to develop clinical prognostic models (CPMs) to identify those most likely to die among community-dwelling children under 5 years presenting to care for acute diarrhea. CPMs are algorithms that aid clinicians in interpreting clinical findings and making clinical decisions[8]. Building on this body of literature, we used machine learning methods on data from two large multi-center studies to derive and validate prediction models for death, both during treatment and post-discharge from treatment, with the hopes of reliably identifying children that would most benefit from intervention.

## METHODS

### Study Population for Derivation Cohort 1 (GEMS)

We derived CPMs for death using data from cases from The Global Enteric Multicenter Study (GEMS), which has previously been described in-depth[7, 9]. GEMS was a prospective case-control study of acute moderate to severe diarrhea (MSD) in children 0-59 months of age in 7 sites in Africa and Asia (Mali, The Gambia, Kenya, Mozambique, Bangladesh, India, and Pakistan). Data were collected in December 2007 – March 2011. MSD was defined as 3 or more looser than normal stools in the previous 24 hours lasting 7 days or less, and had to be new-onset (after ≥7 days diarrhea-free) accompanied by one or more of the following: dysentery (blood in stool observed by the caretaker, clinician, or laboratory), dehydration (decreased skin turgor, sunken eyes more than normal, or IV rehydration prescribed or given), or hospital admission. MSD cases were enrolled at initial presentation to a sentinel health center or hospital serving the site’s censused population. Participants received care consistent with WHO guidelines, including antibiotic treatment for dysentery and suspected cholera, zinc therapy, and nutritional support for children with severe acute malnutrition. Using standardized questionnaires, demographics, epidemiological, and clinical information was collected at presentation from caregivers. In addition, clinic staff conducted physical exams, including anthropometry and stool sample collection which have undergone conventional and molecular testing to ascertain diarrhea etiology. After approximately 60 (up to 91) days after enrollment, fieldworkers visited the homes of participants to repeat anthropometry and collect standardized clinical and epidemiological information.

Participants’ caregivers provided informed consent, in writing or witnessed if caregivers were illiterate. The GEMS study protocol was approved by ethical review boards at each field site and the University of Maryland, Baltimore, USA.

### Study Population for Validation Cohort (Kilifi)

We externally validated our CPMs using data from the Kilifi Health and Demographic Surveillance System (KHDSS) and Kilifi County Hospital (KCH) in Kenya[2]. Children 2-59 months of age who presented with diarrhea to KCH and were resident within the KHDSS were enrolled between January 2007 and December 2015. Similar systematic demographic, epidemiological, and clinical information was collected at admission to KCH. Diarrhea was defined as 3 or more looser than normal stools in the previous 24 hours. Inpatient treatment was provided as per WHO guidelines, including treatment for severe acute malnutrition. Their survival status after hospital discharge was followed through quarterly census in the KHDSS up to August 2017.

Participants’ caregivers provided written informed consent. The study was approved by the Kenya Medical Research Institute (KEMRI) National Ethical Review Committee.

### Outcomes

We examined three related outcomes: death at any time after enrollment at the health facility (after admission to the health center), death at the enrolment health center (after admission, before discharge), and post-discharge death (reported by caregiver (GEMS) or census (KHDSS) reported death after being discharged from medical care within 91 days post-enrollment). Children who died at the enrolment health center or for whom post-discharge follow-up data were missing were excluded from the post-discharge death analysis.

### Predictive Variables

Over 130 potential GEMS predictors were considered, including descriptors of the child, household, and community (Supplemental Table S1). We did not consider aggregate scores as potential predictors (e.g. wealth index), as their clinical use would necessitate collecting multiple variables, each of which were already individually considered in the CPM.

### Statistical Analysis

We developed our CPMs using a multistep process. First, we screened variables using random forests to rank possible predictors by their predictive importance. Random forests are an ensemble learning method whereby multiple decision trees (1000 throughout this analysis) are built on bootstrapped samples of the data with only a random sample of potential predictors considered at each split, thereby decorrelating the trees and reducing variability[10]. In this analysis, the number of variables considered at each split was equal to the square root of the total number of potential variables, rounded down. We defined predictive importance as the reduction in mean squared prediction error that would be achieved by including the variable in the predictive model on out-of-bag samples (i.e. observations not in the bootstrapped sample).

Second, we used repeated cross-validation to assess generalizable model discrimination. For each of 100 iterations, separate logistic regression and random forest regression models were fit to a random 80% sample of the full analytic dataset (training set) using a subset of the top-ranked predictive variables. We examined the top 1-10, 15, 20, 30, 40, and 50 of predictors. Each of these models were then used to predict the outcome on the remaining 20% of the analytic data (testing set). We used the receiver operating characteristic (ROC) curves and the cross-validated C-statistic (area under the ROC curve (AUC)) to assess model discrimination. Discrimination is defined as how well a model can separate individuals who will or will not experience the outcome, in this case death.

Third, we assessed model calibration, or agreement between the predicted and observed risk of the outcome. We assessed calibration-in-the-large, or calibration intercept, by using logistic regression to estimate the mean while subtracting out the estimate (model the log-odds of the true status, offset by the CPM-predicted log-odds). We assessed the spread of the estimated probabilities using calibration slope. To do this, we fit a logistic regression model CPM-predicted log-odds as the independent variable and log-odds of the true status as the dependent variable. We also graphically assessed “moderate calibration.” We calculated the predicted probability of death for each child in a given analysis using each iteration of each n-variable model fit. We then binned these predicted probabilities into deciles, and calculated the proportion of each decile who truly experienced the outcome for each iteration of each n-variable model. We then calculated the mean predicted probability and mean observed proportion for each decile across iterations, and then plotted these averages for each n-variable model[11] (see GitHub).

### Sensitivity Analyses

We undertook a variety of sensitivity and subgroup analyses in the GEMS data to validate our predictive models. First, we explored age-strata specific CPMs for children 0-11months, 12-23months, and 24-59 months. Second, we explored site-specific CPMs. Finally, we fit a model to one continent and validated it on the other as a quasi-external validation. All analysis was conducted in R 4.0.2 using the packages “ranger,” “cvAUC,” and “pROC.”

### External Validation and Comparison to Known Risk Factors

We fit our final CPM in GEMS data, and then applied it to the Kilifi data to evaluate its performance in a new population. As a sensitivity analysis, we fit our CPM to GEMS data only from Kenya, and evaluated its performance in Kilifi data. As an additional evaluation, we assessed how our CPM would have performed as a screening test to identify children at highest risk of dying after presenting to care. We evaluated this by using the final CPM to calculate the predicted probability of death for children in GEMS. We then explored test performance (sensitivity, specificity, etc.) of different predicted probability cutoffs. Previous studies have identified age and MUAC as key risk factors for death following diarrhea[12]. Given the variables identified as top predictors (see Results), we also compared the performance of our CPM as a screening tool to specific patient subpopulations known to be at elevated mortality risk, namely children 0-6 months of age, and children with MUAC<12.5.

## RESULTS

### Death in children following acute diarrhea in GEMS

There were 9439 children with MSD enrolled in GEMS. Of these, 840 children were excluded for having missing follow-up data, and 79 were excluded for having follow-up data outside of the 91 day study follow-up period, leaving an analytic sample of 8520. An additional 460 observations were dropped for missing predictor data, leaving 8060. Of these, 165 (2.0%) of children died by 91 days after enrollment, including 43 (0.5%) during treatment, and 122 (1.5% of remaining) after discharge (Supplemental Figure S1).

### Derivation of a CPM to identify children likely to die following acute diarrhea using GEMS data

In GEMS data, the top predictors of death after enrollment are listed in Table 1 and were: mid-upper arm circumference (MUAC), respiratory rate, temperature, age (months), number of people living in the household, number of days of diarrhea at presentation, how much the child has been offered to drink since diarrhea began, number of children <60 months old living in the household, abnormal hair (e.g. sparse, loose, straight, etc.), and number of rooms used for sleeping, with an AUC of 0.83 (95% CI: 0.81, 0.86) for a 10-variable model. The logistic regression models consistently performed better than the random forest regressions (see Supplemental Figure S2), so we present only the logistic regression models moving forward.

The maximum AUC attained with the model predicting any death after enrollment was 0.88 (95% CI: 0.87, 0.90) with a model of 30 variables, while an AUC of 0.84 (95% CI: 0.82, 0.86), 0.86 (95% CI: 0.84, 0.88) and 0.86 (95% CI: 0.84, 0.88) was obtained with a CPM of 2, 5, and 10 variables, respectively (Supplemental Figure S2). At a sensitivity of 0.80, we achieved a specificity of 0.75 with 10 predictors, and at a sensitivity of 0.90, a specificity of 0.62 (Figure 1). For the CPM predicting any death after enrollment, the calibration-in-the-large, or intercept, was consistently close to 0 for models with 1 to 10 predictor variables, indicating the predicted probability of death was close to the average observed probability of death. The calibration slope was consistently close to 1, indicating the spread of predicated probabilities of death was similar to the spread of observed probabilities for models including 1 to 10 predictor variables (Table 2, Figure 2). The CPM to predict any death (AUC=0.86, 95% CI: 0.84, 0.88) had very similar discriminative ability compared to the model only predicting death in treatment (AUC=0.85, 95% CI: 0.82, 0.88) and death post-discharge (AUC=0.86, 95% CI: 0.84, 0.88). Top predictors were similar across all three models. Odds ratios for the 10-variable model predicting any death are shown in Supplemental Table S2.

### External validation of a CPM to identify children likely to die following acute diarrhea

Given the discriminative performance observed in Table 1 and Figure S2, we elected to proceed with a single CPM for death after enrollment, with MUAC and respiratory rate as predictors. The CPM had good performance on internal cross-validation in GEMS (AUC=0.85, 95% CI: 0.82, 0.88), with a decrease in discriminative ability at external validation in Kilifi data (AUC=0.74, 95% CI: 0.71, 0.77). On average, the CPM underestimated the probability of death (calibration intercept=0.82, 95% CI:0.68, 0.97), and predictions tended to be too extreme (calibration slope=0.61, 95% CI: 0.52, 0.70) (Table 2, Figure 2). Model performance was similar when the CPM was derived only in GEMS data from Kenya and validated on data from Kilifi (see Supplemental Figure S3 and Table S3).

**Table 1:** **DEATH:** Variable importance ordering and cross-validated average overall AUC, AUC by timing of death, and 95% confidence intervals for a 5 (bold) and 10 (italicized) variable logistic regression model for predicting death 60/90 days after acute diarrhea presentation (enrollment) in children 0-59mo in 7 LMICs derived from GEMS data.

|  | Death at any time | Death in treatment | Death after discharge |
| --- | --- | --- | --- |
| Variable/<br>Patient<br>Subset | n=165/8060 | 43/8060 | 122/8017 |
| 1 | MUAC | MUAC | MUAC |
| 2 | Respiratory rate | Respiratory rate | Respiratory rate |
| 3 | Temperature | Age (months) | Temperature |
| 4 | Age (months) | Temperature | Age (months) |
| 5 | Num. people living in household | Num. people living in household | Num. people living in household |
| 6 | Num. days of diarrhea at presentation | Num. days of diarrhea at presentation | Num. days of diarrhea at presentation |
| 7 | Since diarrhea starts, how much offering child to drink | Num. children <60months live in household | Since diarrhea starts, how much offering child to drink |
| 8 | Num. children <60months live in household | Dad_live | Num. children <60months live in household |
| 9 | Abnormal hair (e.g. sparse, loose, straight) | Num. rooms used for sleeping | Num. rooms used for sleeping |
| 10 | Num. rooms used for sleeping | Since diarrhea starts, how much offering child to drink | Abnormal hair (e.g. sparse, loose, straight) |
| AUCs | **0.86 (0.84, 0.88)** | **0.87 (0.84, 0.89)** | **0.85 (0.83, 0.87)** |
|  | *0.86 (0.84, 0.88)* | *0.85 (0.82, 0.88)* | *0.86 (0.84, 0.88)* |

**Table 2:** Calibration intercept and slope

| Number of predictor variables | GEMS 0-59mo Intercept (95% CI) | Slope (95% CI) | GEMS-derived model applied to KILIFI data Intercept (95% CI) | Slope (95% CI) |
| --- | --- | --- | --- | --- |
| 1 | $3.6 \times 10^{-2}$ ($-3.4 \times 10^{-1}$, $3.8 \times 10^{-1}$) | 1.0 (0.75, 1.3) | | |
| 2 | $3.5 \times 10^{-2}$ ($-3.4 \times 10^{-1}$, $3.8 \times 10^{-1}$) | 1.0 (0.76, 1.3) | 0.82 (0.68, 0.97) | 0.61 (0.52, 0.70) |
| 3 | $2.7 \times 10^{-2}$ ($-3.5 \times 10^{-1}$, $3.7 \times 10^{-1}$) | 1.0 (0.77, 1.3) | | |
| 4 | $2.4 \times 10^{-2}$ ($-3.6 \times 10^{-1}$, $3.7 \times 10^{-1}$) | 1.0 (0.79, 1.3) | | |
| 5 | $2.1 \times 10^{-2}$ ($-3.6 \times 10^{-1}$, $3.7 \times 10^{-1}$) | 1.0 (0.78, 1.3) | | |
| 6 | $1.7 \times 10^{-2}$ ($-3.7 \times 10^{-1}$, $3.6 \times 10^{-1}$) | 0.99 (0.75, 1.2) | | |
| 7 | $2.7 \times 10^{-3}$ ($-3.8 \times 10^{-1}$, $3.5 \times 10^{-1}$) | 0.95 (0.73, 1.2) | | |
| 8 | $1.1 \times 10^{-2}$ ($-3.8 \times 10^{-1}$, $3.6 \times 10^{-1}$) | 0.95 (0.73, 1.2) | | |
| 9 | $1.8 \times 10^{-2}$ ($-3.7 \times 10^{-1}$, $3.7 \times 10^{-1}$) | 0.94 (0.72, 1.2) | | |
| 10 | $2.5 \times 10^{-2}$ ($-3.6 \times 10^{-1}$, $3.8 \times 10^{-1}$) | 0.93 (0.71, 1.2) | | |

**Figure 1:**
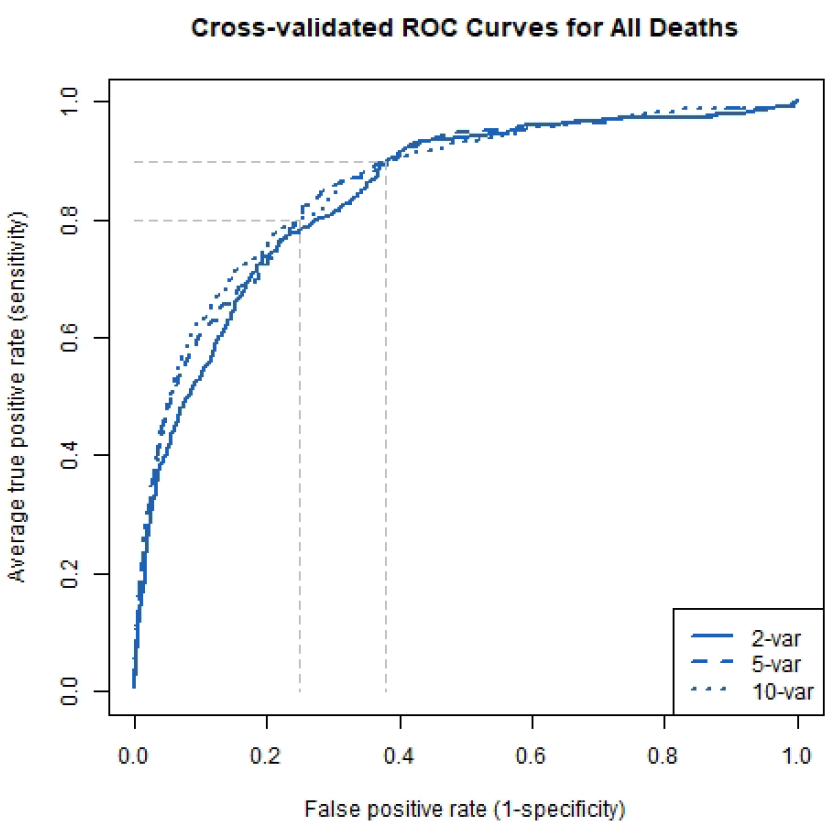
ROC curves. Average ROC curves from the cross-validated logistic regression models predicting growth faltering and death with 2, 5, and 10 predictors. The faded dashed lines represent examples of specificities (1-false positive rate) that could be achieved with a sensitivity (true positive rate) of 0.80 for prediction of each outcome.

**Figure 2:**
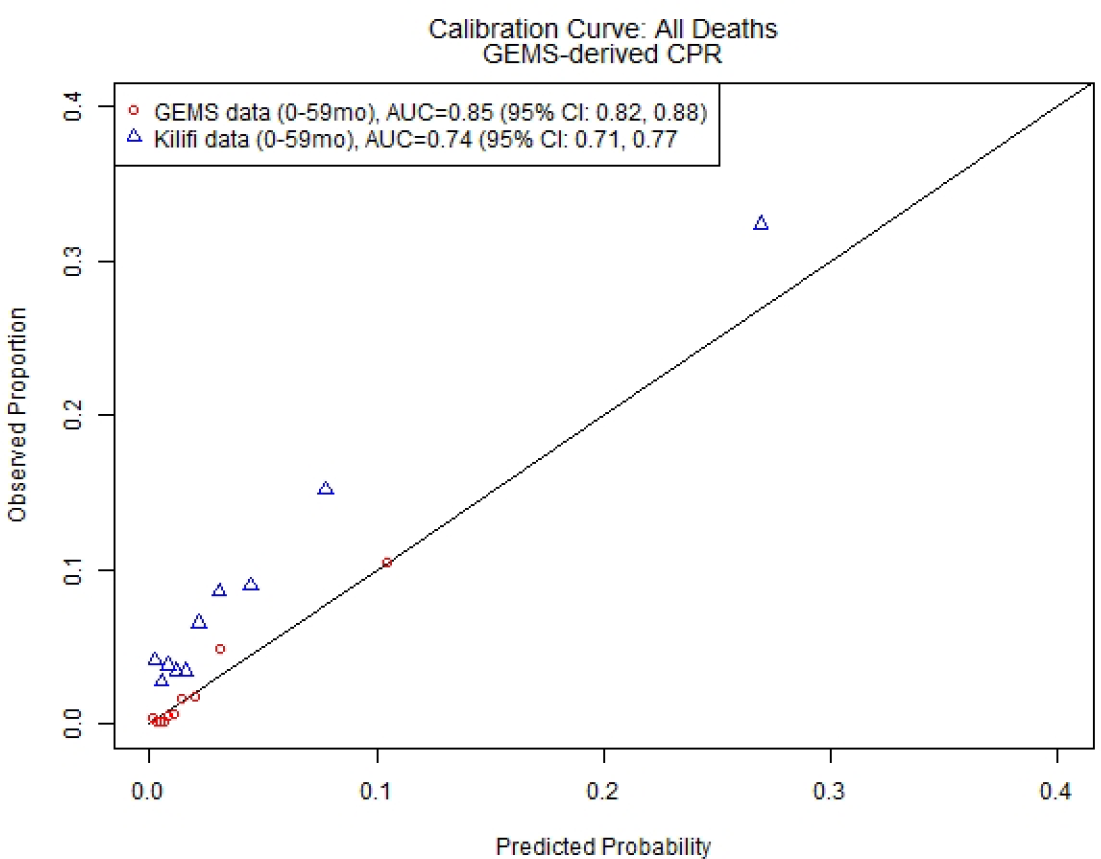
2-Variable CPM for death after presentation. Calibration curve and discriminative ability of 2-variable (MUAC, respiratory rate) model predicting death after presentation to care for acute diarrhea in LMICs.

### Discriminative performance of the CPM to identify children likely to die following acute diarrhea was generally consistent across age and location subpopulations

The results of the sensitivity analyses are presented in Supplemental Table S4. Top predictor variables were highly consistent across models and included patient demographics, patient symptoms, and indicators of household wealth. While the CPM fit to patients age 24-59 months had a slightly higher AUC compared to the overall model (AUC=0.91, 95% CI: 0.87, 0.95 for 2-variables for ages 24-59months vs AUC=0.84, 95% CI: 0.82, 0.86), this is the patient population with the lowest overall risk of death (Supplemental Table S5). The CPMs fit solely to each of the GEMS sites in Africa all had similar performance to the overall model, whereas there were too few outcomes in the GEMS sites in Asia to fit country-specific models (see Supplemental Tables S4 and S6). In our quasi-external validation, the model was fit to GEMS data from all the sites in Africa, the AUC was almost identical to the overall model, and performed excellently in GEMS data from the Asian sites (AUC=0.93, 95% CI 0.90, 0.96) (see Supplemental Table S4).

### A screening tool based on our CPM could improve upon risk-factor based screening to identify children likely to die following acute diarrhea

Using the 2-variable CPM derived in GEMS described above, we explored how accurately our CPM identified children who went on to die during our study period. Using a CPM-derived predicted probability of ≥0.10 as a positive screen for risk of death, we observed a sensitivity (Se) of 0.28 and a negative predictive value (NPV) of 0.98 in GEMS. In contrast, using an observed MUAC of <12.5 as a positive screen for death resulted in a Se of 0.66 and a NPV of 0.99. However, almost 6 times as many children screened positive for risk of death using the MUAC-based approach compared to our CPM-based approach (17.5% vs 3.1% of patients screen positive). Increasing the predicted probability threshold of our CPM screen led to decreasing sensitivity and fewer children screening positive (see Supplemental Table S7).

## DISCUSSION

We used a combination of machine learning and conventional regression methods to derive and externally validate clinical prognostic models for death following acute diarrhea. Our CPM to predict death in community-dwelling children at the time they present for care for acute MSD had good discriminative ability in the derivation dataset (GEMS AUC=0.84, 95% CI: 0.82, 0.86) as well as at external validation (Kilifi AUC=0.74, 95% CI: 0.71, 0.77). There have previously been limited efforts to identify which children are more likely to die after presenting to care for acute diarrhea. While a number of studies have explored risk factors of post-discharge mortality after seeking care for diarrhea in LMICs [1, 2, 13], prediction tools have been lacking. Our CPM for mortality suggests the potential for parsimonious clinical prognostic model(s) to guide appropriate triage and follow-up for young children with acute diarrhea.

In our model derivation, we found a similar set of top predictors for the categories of: any death after enrollment, death during treatment, and death after discharge, as well as for different age subgroups and GEMS study sites. Mid upper arm circumference (MUAC) was the top predictor for all CPMs. Low MUAC has previously been recognized as a good predictor of mortality[14, 15]. While MUAC is somewhat affected by acute dehydration, it is much less impacted than other markers of malnutrition (e.g. weight-for-length z-score)[16, 17]. In addition, MUAC is currently only recommended as an indicator of SAM in children 6 months of age and above, there is growing evidence in support of its use in children <6 months[18-20]. The use of MUAC as a key predictor in risk of death is also supported by a recent prospective cohort study (CHAIN) of children 2-23 months of age who presented to care for acute illness in 6 LMICs. The authors found that nutritional status was directly associated with death 30 days from admission, capturing a range of underlying risks[5], and that MUAC was a top predictor of death[21].

We found similar discriminative ability for predicting death at different time points (in treatment, post-discharge), in different age subsets, and at different GEMS study sites. As predictors in our derivation dataset were collected only at enrollment, we were unable to examine if updated values at discharge could improve our post-discharge mortality prediction. However, others have found no differences in cross-validated discriminative ability even with updated predictors at discharge[5].

Our CPM is promising as a screening tool to identify children likely to die after presentation, and therefore who may benefit from more intensive care and follow-up. While young age and poor nutritional status are known risk factors of poor diarrhea outcomes, our CPM performed better than simple age and MUAC based cutoff screening criteria. The highest screening sensitivity was achieved by using MUAC<12.5 as a screening cutoff for all children age 0-59mo (Se=0.66), meaning this screening protocol correctly captured two-thirds of children who died in the study period. Such screening at presentation would allow for early intervention and intensive follow-up to potentially avoid these deaths. However, using these criteria, 17.5% of presenting children would have screened positive. This may be an unrealistically large portion of children for whom to provide intensive treatment and follow-up care. In contrast, if our CPM-calculated predicted probability of death was used as a screening tool in GEMS, we would have correctly identified 28% of children who went on to die within the study period, while only having 3.1% of those presenting to care screen positive. Future research should explore the effectiveness, viability, and ethics of using such predictive screening tools to allocate limited resource-intensive acute and follow-up care.

Utilization of a CPM at time of clinical presentation could offer a timely and efficient way to identify children most likely to benefit from targeted, resource-intensive interventions such as additional staffing during treatment, extended treatment at care facility, and at-home follow-up care post-discharge. However, this assumes the children predicted to die would avoid death with adequate intervention. A recent systematic review of interventions for post-acute diarrhea sequelae suggests that the majority of existing intervention strategies are not effective at reducing mortality[22]. While nutritional status was a top predictor of mortality in our study, it is important to note that all cases of severe acute malnutrition were treated according to WHO guidelines. This suggests that treating malnutrition at presentation is insufficient to avert the mortality seen in our study. Similarly, all cases of dysentery were treated with antibiotics according to WHO guidelines, but previous research has shown that *Shigella* spp was common among children without dysentery (and therefore did not meet antibiotic recommendations) who died in GEMS[23]. Additional research is needed in this area to develop effective interventions that reduce longer-term mortality risk following acute diarrhea in young children.

Our study has a number of strengths and limitations. We derived CPMs for death from a multi-site, prospective study that included longitudinal follow-up with extensive etiologic testing. We used random forests for variable selection, which do not require assumptions to be made about the underlying variables. They also tend to outperform[24] conventional model building techniques. Our modeling strategy required complete predictor data, and we dropped approximately 5% of eligible observations in GEMS due to missing predictor data. Fortunately, the distribution of top predictive variables was generally very similar between dropped and retained observations (Supplemental Table S8). We were also able to externally validate our CPM in a similar setting with a similar distribution of patient characteristics (Supplemental Figure S4), with promising results. However, the patient population was likely less healthy at presentation in our external validation dataset than our derivation datasets, as evidence by the higher mortality rate (Supplemental Table S6). While the CPM had good discriminative ability at external validation, the model calibration needs improvement and should be prospectively validated before its ready for clinical application.

In conclusion, we used data from a large multi-country study to derive and an external dataset to validate clinical prognostic models for death. Our findings suggest it is possible to identify children most likely to die after presenting to care for acute diarrhea. This could represent a novel and cost-effective way to target resources for the prevention of childhood mortality.

## Supporting information

DeathONLYCPR_suppl

## Data Availability

GEMS data is available from clinepidb.org
Kilifi data is available from the authors upon request and approval.

https://clinepidb.org/ce/app/workspace/analyses/DS_841a9f5259/new/details

