## Supplementary material for "Derivation and external validation of a clinical prognostic model identifying children at risk of death following presentation for diarrheal care": DeathONLYCPR_suppl

### SUPPLEMENT

Table S1: Full list of considered predictor variables

Study site (site)

Child sex (f3_gender)

Loss of skin turgor (f3_drh_turgor)

Intravenous rehydration (f3_drh_iv)

Hospitalized (f3_drh_hosp)

Your relationship to the child (f4a_relationship)

Where child’s father lives (f4a_dad_live)

Primary caregiver’s max school (f4a_prim_schl)

People living in house 6 months (f4a_ppl_house)

Children under 60 months in the house (f4a_yng_children)

How many rooms used for sleeping (f4a_slp_rooms)

Predominant floor (f4a_floor)

Electricity (f4a_house_elec)

Bicycle/rickshaw (f4a_house_bike)

Telephone (f4a_house_phone)

Television (f4a_house_tele)

Car/truck (f4a_house_car)

Animal-drawn cart (f4a_house_cart)

Motorcycle/scooter (f4a_house_scoot)

Refrigerator (f4a_house_fridge)

Agricultural land (f4a_house_agland)

Radio (f4a_house_radio)

Boat with motor (f4a_house_boat)

None of the above assets (f4a_house_none)

Electricity (f4a_fuel_elec)

Biogas (f4a_fuel_biogas)

Straw/shrubs/grass (f4a_fuel_grass)

Liquid propane gas (f4a_fuel_propane)

Coal/lignite (f4a_fuel_coal)

Animal dung (f4a_fuel_dung)

Natural gas (f4a_fuel_natgas)

Charcoal (f4a_fuel_charcoal)

Agricultural crop residue (f4a_fuel_crop)

Kerosene (f4a_fuel_kero)

Wood (f4a_fuel_wood)

Other fuel (f4a_fuel_other)

Goat (f4a_ani_goat)

Sheep (f4a_ani_sheep)

Dog (f4a_ani_dog)

Cat (f4a_ani_cat)

Cow (f4a_ani_cow)

Rodents (f4a_ani_rodents)

Fowl (f4a_ani_fowl)

Other animal (f4a_ani_other)

No animals (f4a_ani_no)

Water piped to house (f4a_water_house)

Covered well in house/yard (f4a_water_covwell)

Water piped into yard (f4a_water_yard)

Covered public well (f4a_water_covpwell)

Public tap (f4a_water_pubtap)

Protected spring (f4a_water_prospring)

Open well in house/yard (f4a_water_well)

Unprotected spring (f4a_water_unspring)

Open public well (f4a_water_pubwell)

River/stream (f4a_water_river)

Pond/lake (f4a_water_pond)

Deep tube well (f4a_water_deepwell)

Rainwater (f4a_water_rain)

Shallow tube well (f4a_water_shallwell)

Bought water (f4a_water_bought)

Other water source (f4a_water_othr)

Bore hole (f4a_water_bore)

Main source of drinking water (f4a_ms_water*)

How often is water available (f4a_water_avail)

Did you give the child stored water (f4a_store_water)

Do you usually treat drinking water? (f4a_trt_water)

Usual treatment method (f4a_trt_method)

How are child’s feces disposed (f4a_disp_feces)

Facility used to dispose of feces (f4a_fac_waste)

How many households share facility? (f4a_share_fac)

Wash hands before eating? (f4a_wash_eat)

Wash hands before cooking (f4a_wash_cook)

Wash hands before you nurse? (f4a_wash_nurse)

Wash hands after you defecate (f4a_wash_def)

Wash hands after handling animals (f4a_wash_animal)

Wash hands after cleaning a child (f4a_wash_child)

Wash hands other times (f4a_wash_othr)

What do you use to wash your hands? (f4a_wash_use)

Is the child currently breastfed? (f4a_breastfed)

How long as this diarrhea episode lasted (days)? (f4a_drh_days)

Maximum number of loose stools (f4a_max_stools)

Blood in stools (f4a_drh_blood)

Vomiting 3 or more times per day (f4a_drh_vomit)

Very thirsty (f4a_drh_thirst)

Drank much less than usual (f4a_drh_lessdrink)

Belly pain (f4a_drh_bellypain)

Irritable or restless (f4a_drh_restless)

Decreased activity or lethargy (f4a_drh_lethrgy)

Loss of consciousness (f4a_drh_consc)

Rectal straining (f4a_drh_strain)

Rectal prolapse (f4a_drh_prolapse)

Cough (f4a_drh_cough)

Convulsions (f4a_drh_conv)

Very thirsty (f4a_cur_thirsty)

Wrinkled skin (f4a_cur_skin)

Irritable or restless (f4a_cur_restless)

Dry mouth (f4a_cur_drymouth)

Fast breathing (f4a_cur_fastbreath)

ORALITE or ORS (f4a_hometrt_ors)

Homemade fluid (f4a_hometrt_maize)

Special mile or infant formula (f4a_hometrt_milk)

Home remedy/herbal medication (f4a_hometrt_herb)

Zinc (f4a_hometrt_zinc)

No special remedies given (f4a_hometrt_none)

Any other liquids (f4a_hometrt_othrliq)

Antibiotics (f4a_hometrt_ab)

Other treatment (f4a_hometrt_othr1)

Other treatment (f4a_hometrt_othr2)

How much offered to drink (f4a_offr_drink)

Seek outside care (f4a_seek_outside)

Pharmacy (f4a_seek_pharm)

Friend/relative (f4a_seek_friend)

Traditional healer (f4a_seek_healer)

Unlicensed practitioner (f4a_seek_doc)

Licensed practitioner (f4a_seek_privdoc)

Bought a remedy (f4a_seek_remdy)

Other hospital/center (f4a_seek_other)

Mid-upper arm circumference (f4b_muac)

Axillary temperature (f4b_temp)

Respiratory rate per minute (f4b_resp)

Chest indrawing (f4b_chest_indrw)

Eyes (f4b_eyes)

Mouth (f4b_mouth)

Skin pinch (f4b_skin)

Mental status (f4b_mental)

Rectal prolapse (f4b_rectal)

Bipedal edema (f4b_bipedal)

Abnormal hair (f4b_abn_hair)

Undernutrition (f4b_under_nutr)

Skin as ‘flaky paint’ appearance (f4b_skin_flaky)

Receive rehydration here (f4b_recommend)

Child was admitted to hospital (f4b_admit)

Child age (months) (base_age)

*f4a_ms_water was recategorized into the following: surface, other unimproved, other improved, piped, other[18, 19]

Figure S1: Flow diagram of study inclusion

GEMS

9439 children enrolled with acute diarrhea (≤7 days duration)

840 missing follow-up data

79 had follow-up data outside of follow-up study period

8520 eligible

460 missing predictor data

8060 included in analysis

- 43 died during treatment

- 122 died after discharge

Figure S2: Number of variables and AUC for random forest regression and logistic regression

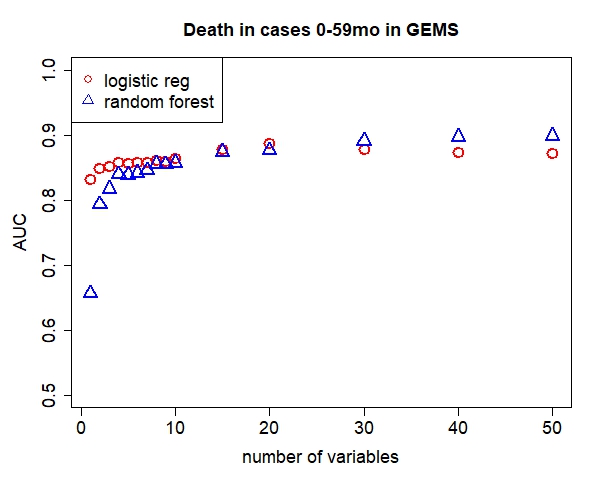

Table S2: Variable importance ordering, cross-validated average AUC, odds ratios, and 95% confidence intervals for logistic regression models predicting death in children 0-59mo in LMICs in GEMS ranked from most to less predictive (highest to lower variance reduction)

| GEMS |  |
| --- | --- |
| AUC (95% CI): 0.86 (0.84, 0.88) |  |
| Variables | OR (95% CI) |
| MUAC | 0.48 (0.43, 0.54) |
| Respiratory rate | 1.03 (1.01 1,04) |
| Temperature | 1.51 (1.28, 1.78) |
| Age (months) | 1.02 (1.00, 1.03) |
| Num. ppl living in household | 1.00 (0.97, 1.02) |
| Num. days of diarrhea at presentation | 1.07 (0.95, 1.21) |
| Since diarrhea starts, how much offering child to drink | 1.35 (1.16, 1.57) |
| Num. children <60months live in household | 0.98 (0.88, 1.09) |
| Abnormal hair (sparse, loose, straight) | 4.03 (2.61, 6.14) |
| Num. rooms in house used for sleeping | 1.02 (0.96, 1.09) |

### Figure S3 Fit model just to GEMS Kenya, see how performs in Kilifi

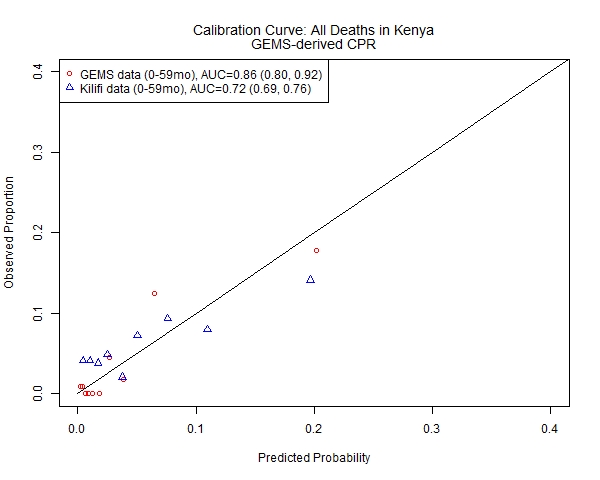

Table S3 Calibration assessment:

| Kenya only GEMS-derived model applied to KILIFI data  Intercept (95% CI) | Slope (95% CI) |
| --- | --- |
| -0.25 (-0.40, -0.10) | 0.50 (0.42, 0.58) |

Supplementary Table S4: **DEATH AT ANY TIME** Variable importance ordering and cross-validated average overall AUC and AUC by patient subset and 95% confidence intervals for a 2 (plain text), 5 (bold), and 10 (italicized) variable logistic regression model for predicting any death in children in GEMS

| Patient Subset | 0-59mo (main text model) | 0-11mo | 12-23mo | 24-59mo |
| --- | --- | --- | --- | --- |
|  | 0.84 (0.82, 0.86) | 0.80 (0.77, 0.83) | 0.80 (0.76, 0.85) | 0.91 (0.87, 0.95) |
| AUCs | **0.86 (0.84, 0.88)** | **0.82 (0.79, 0.85)** | **0.81 (0.77, 0.85)** | **0.93 (0.89, 0.96)** |
|  | *0.86 (0.84, 0.88)* | *0.82 (0.79, 0.85)* | *0.85 (0.81, 0.88)* | *0.90 (0.86, 0.95)* |
| 1 | MUAC | MUAC | MUAC | MUAC |
| 2 | Respiratory rate | Temperature | Temperature | Abnormal hair (e.g. sparse, loose, straight) |
| 3 | Temperature | Respiratory rate | Num. people living in household | Skin has “flaky paint” appearance |
| 4 | Age (months) | Age (months) | Respiratory rate | Respiratory rate |
| 5 | Num. people living in household | Num. people living in household | Num. children <60months live in household | Temperature |
| 6 | Num. days of diarrhea at presentation | Num. days of diarrhea at presentation | Num. days of diarrhea at presentation | Age (months) |
| 7 | Since diarrhea starts, how much offering child to drink | Chest indrawing | Age (months) | Num. days of diarrhea at presentation |
| 8 | Num. children <60months live in household | Where child’s father lives | Abnormal hair (e.g. sparse, loose, straight) | Undernutrition |
| 9 | Abnormal hair (e.g. sparse, loose, straight) | Since diarrhea starts, how much offering child to drink | Num. rooms used for sleeping | Since diarrhea starts, how much offering child to drink |
| 10 | Num. rooms used for sleeping | Num. children <60months live in household | Homemade remedy / herbal medication | Skin pinch |

|  | The Gambia | Mali | Mozambique | Kenya | India | Bangladesh | Pakistan | Fit in data from Africa | Fit in data from Asia |
| --- | --- | --- | --- | --- | --- | --- | --- | --- | --- |
| AUCs |  |  |  |  | too few outcomes, | too few outcomes, | too few outcomes, | 0.83 (0.82, 0.84) | too few outcomes, |
|  | **0.78 (0.77, 0.80)** | **0.88 (0.87, 0.89)** | **0.79 (0.77, 0.80)** | **0.88 (0.85, 0.91)** | model does not converge | model does not converge | model does not converge | **0.84 (0.83, 0.85)** | model does not converge |
|  | *0.78 (0.76, 0.80)* | *0.87 (0.85, 0.88)* | *0.77 (0.75, 0.79)* | *0.84 (0.81, 0.87)* |  |  |  | *0.83 (0.82, 0.84)* |  |
| 1 | MUAC | MUAC | MUAC | MUAC |  |  |  | MUAC |  |
| 2 | Respiratory rate | Temperature | Temperature | Age (months) |  |  |  | Age (months) |  |
| 3 | Age (months) | Num. people living in household | Age (months) | Respiratory rate |  |  |  | Respiratory rate |  |
| 4 | Num. days of diarrhea at presentation | Respiratory rate | Respiratory rate | Temperature |  |  |  | Temperature |  |
| 5 | Num. people living in household | Age (months) | Undernutrition | Undernutrition |  |  |  | Num. people living in household |  |
| 6 | Temperature | Chest indrawing | Abnormal hair (e.g. sparse, loose, straight) | Since diarrhea starts, how much offering child to drink |  |  |  | Num. days of diarrhea at presentation |  |
| 7 | Num. rooms used for sleeping | Num. rooms used for sleeping | Since diarrhea starts, how much offering child to drink | Bipedal edema |  |  |  | Num. children <60months live in household |  |
| 8 | Num. children <60months live in household | How are child’s feces disposed? | Num. people living in household | Num. days of diarrhea at presentation |  |  |  | Since diarrhea starts, how much offering child to drink |  |
| 9 | Mental status | Num. children <60months live in household | Where child’s father lives | Num. people living in household |  |  |  | Undernutrition |  |
| 10 | Chest indrawing | Special milk or infant formula | Num. children <60months live in household | Main source of drinking water |  |  |  | Num. rooms used for sleeping |  |
|  |  |  |  |  |  |  |  | 2-variable CPM performance in data from Asia |  |
|  |  |  |  |  |  |  |  | 0.93 (0.90, 0.96) |  |

Supplementary Table S5: **DEATHS BY AGE**

| GEMS |  |  |  |
| --- | --- | --- | --- |
|  | 0-11mo | 12-23mo | 24-59mo |
| N | 3383 | 2779 | 1898 |
| Any died | 100 (3.0%) | 46 (1.7%) | 19 (1.0%) |
| In treatment | 23 (0.7%) | 12 (0.4%) | 8 (0.4%) |
| After treatment* | 77 (2.3%) | 34 (1.2%) | 11 (0.6%) |
| Kilifi |  |  |  |
| N | 1385 | 989 | 527 |
| Any died | 130 (9.2%) | 76 (8.7%) | 53 (10.1%) |
| In treatment | 70 (5.0%) | 36 (3.6%) | 21 (4.0%) |
| After treatment* | 60 (4.6%) | 40 (4.2%) | 32 (6.3%) |

* Percentages reflect the number of kids who died at home out of all the children discharged alive (did not die during treatment)

Supplementary Table S6: **DEATHS BY SITE**

| GEMS |  |  |  |  |  |  |  |  |
| --- | --- | --- | --- | --- | --- | --- | --- | --- |
|  | Overall | The Gambia | Mali | Mozamb. | Kenya | India | Bdesh | Pakistan |
| N | 8060 | 859 | 1783 | 518 | 1122 | 1473 | 1348 | 957 |
| Any died | 165 (2.5%) | 37 (4.3%) | 23 (1.3%) | 39 (7.5%) | 43 (3.8%) | 2 (0.1%) | 6 (0.4%) | 15 (1.6%) |
| In treatment | 43 (0.5%) | 17 (2.0%) | 3 (0.2%) | 16 (3.1%) | 2 (0.2%) | 0 (0%) | 5 (0.4%) | 0 (0%) |
| After treatment* | 122 (1.5%) | 20 (2.4%) | 20 (1.1%) | 23 (4.6%) | 41 (3.7%) | 2 (0.1%) | 1 (0.1%) | 15 (1.6%) |
| Kilifi |  |  |  |  |  |  |  |  |
| N | 2901 |  |  |  |  |  |  |  |
| Any died | 259 (8.9%) |  |  |  |  |  |  |  |
| In treatment | 127 (4.4%) |  |  |  |  |  |  |  |
| After treatment* | 132 (4.8%) |  |  |  |  |  |  |  |

* Percentages reflect the number of kids who died at home out of all the children discharged alive (did not die during treatment)

Supplementary Table S7: **Test performance** of different screening criteria for identifying children likely to die at any point after presenting to care for acute diarrhea in GEMS data.

|  | **Proportion of patients who screen positive** | **Sensitivity**  **T+\|D+** | **Specificity**  **T-\|D-** | **PPV**  **D+\|T+** | **NPV**  **D-\|T-** |
| --- | --- | --- | --- | --- | --- |
| **Ages 0-6mo** | **17.7%** | **0.30** | **0.83** | **0.03** | **0.98** |
| **Ages >6mo to 59mo** | 82.3% | 0.70 | 0.17 | 0.02 | 0.97 |
| **Ages 0-6mo and MUAC <12.5** | 6.4% | 0.20 | 0.94 | 0.06 | 0.98 |
| **Ages >6mo to 59mo and MUAC<12.5** | 11.2% | 0.46 | 0.90 | 0.08 | 0.99 |
| **Ages 0-59mo and MUAC <12.5** | **17.5%** | **0.66** | **0.83** | **0.08** | **0.99** |
| **CPM* predicted probability ≥0.05** | 8.3% | 0.28 | 0.97 | 0.12 | 0.99 |
| **CPM* predicted probability ≥0.10** | **3.1%** | **0.28** | **0.97** | **0.19** | **0.98** |
| **CPM* predicted probability ≥0.15** | 1.6% | 0.17 | 0.99 | 0.22 | 0.98 |
| **CPM* predicted probability ≥0.20** | 1.2% | 0.13 | 0.99 | 0.23 | 0.98 |

*clinical prognostic model (CPM) including MUAC and respiratory rate, fit a single time to all ages 0-59mo

Supplementary Table S8: **Patient characteristics** of observations included in analysis and dropped due to missing predictor data

|  | **Dropped due to missing predictor data** | **Included in analysis** | **Eligible** |
| --- | --- | --- | --- |
|  | **(N=460)** | **(N=8060)** | **(N=8520)** |
| **number who died** | 14 (3.0%) | 165 (2.0%) | 179 (2.1%) |
| **age (months)** |  |  |  |
| Mean (SD) | 16.9 (13.2) | 16.9 (12.2) | 16.9 (12.3) |
| Median [Min, Max] | 12.0 [0, 59.0] | 14.0 [0, 59.0] | 13.0 [0, 59.0] |
| **site** |  |  |  |
| The Gambia | 22 (4.8%) | 859 (10.7%) | 881 (10.3%) |
| Mali | 3 (0.7%) | 1783 (22.1%) | 1786 (21.0%) |
| Mozambique | 52 (11.3%) | 518 (6.4%) | 570 (6.7%) |
| Kenya | 290 (63.0%) | 1122 (13.9%) | 1412 (16.6%) |
| India | 31 (6.7%) | 1473 (18.3%) | 1504 (17.7%) |
| Bangladesh | 27 (5.9%) | 1348 (16.7%) | 1375 (16.1%) |
| Pakistan | 35 (7.6%) | 957 (11.9%) | 992 (11.6%) |
| **MUAC** |  |  |  |
| Mean (SD) | 13.5 (1.67) | 13.7 (1.51) | 13.7 (1.52) |
| Median [Min, Max] | 13.6 [6.83, 20.0] | 13.7 [6.93, 36.8] | 13.7 [6.83, 36.8] |
| **respiratory rate** |  |  |  |
| Mean (SD) | 39.3 (11.4) | 37.6 (8.93) | 37.7 (9.08) |
| Median [Min, Max] | 37.0 [19.5, 80.0] | 36.5 [13.5, 122] | 37.0 [13.5, 122] |
| Missing | 4 (0.9%) | 0 (0%) | 4 (0.0%) |
| **Temperature (C)** |  |  |  |
| Mean (SD) | 37.4 (1.10) | 37.2 (0.981) | 37.2 (0.988) |
| Median [Min, Max] | 37.1 [35.0, 40.7] | 37.0 [33.0, 41.1] | 37.0 [33.0, 41.1] |
| Missing | 1 (0.2%) | 0 (0%) | 1 (0.0%) |
| **Num. ppl living in household** | |  |  |
| Mean (SD) | 6.87 (6.21) | 11.3 (11.9) | 11.1 (11.7) |
| Median [Min, Max] | 5.00 [2.00, 100] | 7.00 [2.00, 229] | 7.00 [2.00, 229] |
| Missing | 2 (0.4%) | 0 (0%) | 2 (0.0%) |

Supplementary Figure S4: **Patient characteristics** of top predictive variables in GEMS derivation dataset and Kilifi external validation dataset

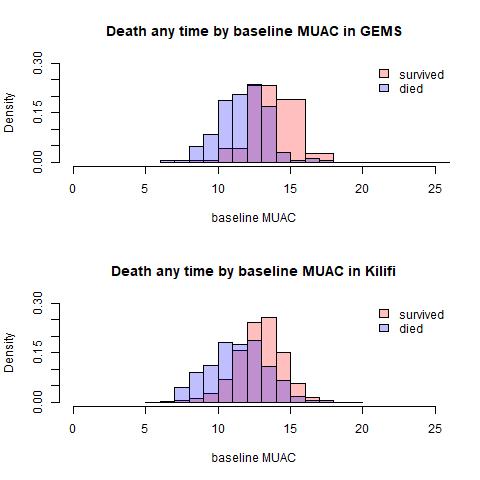

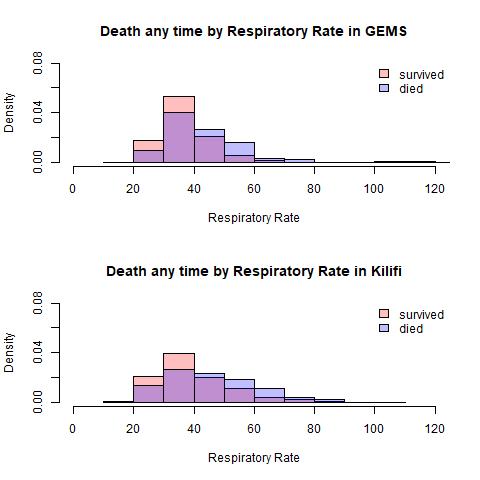

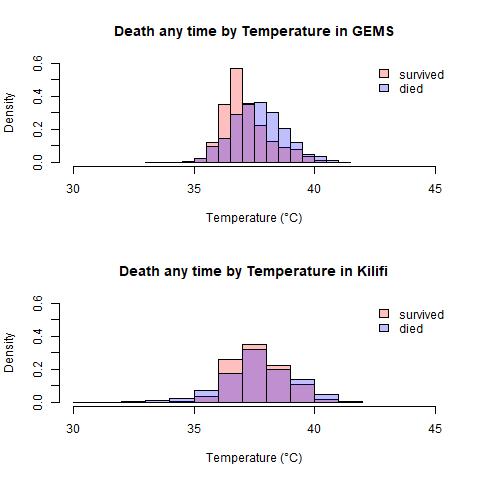

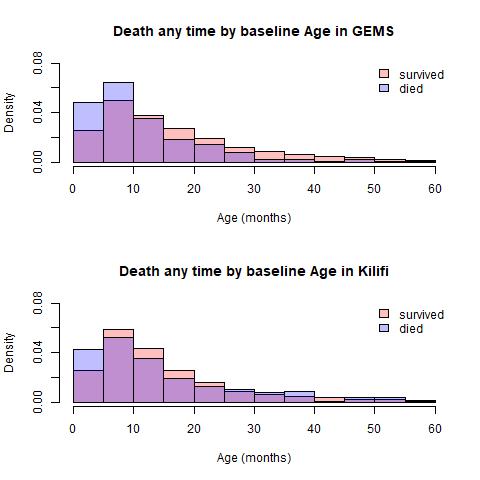

While children 0-6months of age are more likely to die in GEMS than in older age groups, sensitivity of 7-59mo higher since most deaths occur in that larger age category.

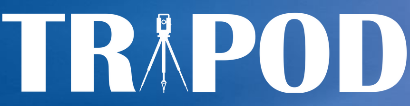
TRIPOD Checklist: Prediction Model Development and Validation

| **Section/Topic** | | **Item** |  |  | **Checklist Item** | **Page** |
| --- | --- | --- | --- | --- | --- | --- |
|  | **Title and abstract** | | | | | |
| Title | | 1 |  | D;V | Identify the study as developing and/or validating a multivariable prediction model, the target population, and the outcome to be predicted. | 1 |
| Abstract | | 2 |  | D;V | Provide a summary of objectives, study design, setting, participants, sample size, predictors, outcome, statistical analysis, results, and conclusions. | 2 |
|  | **Introduction** | | | | | |
| Background and objectives | | 3a |  | D;V | Explain the medical context (including whether diagnostic or prognostic) and rationale for developing or validating the multivariable prediction model, including references to existing models. | 3 |
|  |  | 3b |  | D;V | Specify the objectives, including whether the study describes the development or validation of the model or both. | 3 |
|  | **Methods** | | | | | |
| Source of data | | 4a |  | D;V | Describe the study design or source of data (e.g., randomized trial, cohort, or registry data), separately for the development and validation data sets, if applicable. | 3-5 |
|  |  | 4b |  | D;V | Specify the key study dates, including start of accrual; end of accrual; and, if applicable, end of follow-up. | 3-5 |
| Participants | | 5a |  | D;V | Specify key elements of the study setting (e.g., primary care, secondary care, general population) including number and location of centres. | 3-5 |
|  |  | 5b |  | D;V | Describe eligibility criteria for participants. | 3-5 |
|  |  | 5c |  | D;V | Give details of treatments received, if relevant. | 3-5 |
| Outcome | | 6a |  | D;V | Clearly define the outcome that is predicted by the prediction model, including how and when assessed. | 5 |
|  |  | 6b |  | D;V | Report any actions to blind assessment of the outcome to be predicted. | 3-5 |
| Predictors | | 7a |  | D;V | Clearly define all predictors used in developing or validating the multivariable prediction model, including how and when they were measured. | 5, suppl |
|  |  | 7b |  | D;V | Report any actions to blind assessment of predictors for the outcome and other predictors. | 3-5 |
| Sample size | | 8 |  | D;V | Explain how the study size was arrived at. | 8 |
| Missing data | | 9 |  | D;V | Describe how missing data were handled (e.g., complete-case analysis, single imputation, multiple imputation) with details of any imputation method. | 8 |
| Statistical analysis methods | | 10a |  | D | Describe how predictors were handled in the analyses. | 5-7 |
|  |  | 10b |  | D | Specify type of model, all model-building procedures (including any predictor selection), and method for internal validation. | 5-7 |
|  |  | 10c |  | V | For validation, describe how the predictions were calculated. | 5, suppl |
|  |  | 10d |  | D;V | Specify all measures used to assess model performance and, if relevant, to compare multiple models. | 5-7 |
|  |  | 10e |  | V | Describe any model updating (e.g., recalibration) arising from the validation, if done. | n/a |
| Risk groups | | 11 |  | D;V | Provide details on how risk groups were created, if done. | 7 |
| Development vs. validation | | 12 |  | V | For validation, identify any differences from the development data in setting, eligibility criteria, outcome, and predictors. | 4,5,  13,14 |
|  | **Results** | | | | | |
| Participants | | 13a |  | D;V | Describe the flow of participants through the study, including the number of participants with and without the outcome and, if applicable, a summary of the follow-up time. A diagram may be helpful. | 8, suppl |
|  |  | 13b |  | D;V | Describe the characteristics of the participants (basic demographics, clinical features, available predictors), including the number of participants with missing data for predictors and outcome. | 8, Table S8 |
|  |  | 13c |  | V | For validation, show a comparison with the development data of the distribution of important variables (demographics, predictors and outcome). | Fig S4 |
| Model development | | 14a |  | D | Specify the number of participants and outcome events in each analysis. | 8, Table S6 |
|  |  | 14b |  | D | If done, report the unadjusted association between each candidate predictor and outcome. | n/a |
| Model specification | | 15a |  | D | Present the full prediction model to allow predictions for individuals (i.e., all regression coefficients, and model intercept or baseline survival at a given time point). | Table S2 |
|  |  | 15b |  | D | Explain how to the use the prediction model. | 10, Table S7 |
| Model performance | | 16 |  | D;V | Report performance measures (with CIs) for the prediction model. | Tables 1, S4 |
| Model-updating | | 17 |  | V | If done, report the results from any model updating (i.e., model specification, model performance). | n/a |
|  | **Discussion** | | | | | |
| Limitations | | 18 |  | D;V | Discuss any limitations of the study (such as nonrepresentative sample, few events per predictor, missing data). | 13-14 |
| Interpretation | | 19a |  | V | For validation, discuss the results with reference to performance in the development data, and any other validation data. | 12,13 |
|  |  | 19b |  | D;V | Give an overall interpretation of the results, considering objectives, limitations, results from similar studies, and other relevant evidence. | 11-14 |
| Implications | | 20 |  | D;V | Discuss the potential clinical use of the model and implications for future research. | 12 |
|  | **Other information** | | | | | |
| Supplementary information | | 21 |  | D;V | Provide information about the availability of supplementary resources, such as study protocol, Web calculator, and data sets. | 7 |
| Funding | | 22 |  | D;V | Give the source of funding and the role of the funders for the present study. | 1 |

*Items relevant only to the development of a prediction model are denoted by D, items relating solely to a validation of a prediction model are denoted by V, and items relating to both are denoted D;V. We recommend using the TRIPOD Checklist in conjunction with the TRIPOD Explanation and Elaboration document.
